# COVID-19 Pandemic and Behavioural Response to Self-Medication Practice in Western Uganda

**DOI:** 10.1101/2021.01.02.20248576

**Authors:** Samuel S. Dare, Ejike Daniel Eze, Echoru Isaac, Ibe Michael Usman, Fred Ssempijja, Edmund Eriya Bukenya, Robinson Ssebuufu

**Affiliations:** School of Medicine, Kabale University, Uganda; Faculty of Biomedical Sciences, Kampala International University Western Campus, Uganda; Faculty of Clinical Medicine and Dentistry, Kampala International University Teaching Hospital, Uganda

**Keywords:** Covid-19, self-medication, pandemic, behavioural response, lockdown, drugs

## Abstract

**Background:** Self-medication has become is a serious public health problem globally posing great risks, especially with the increasing number of cases of COVID-19 disease in Uganda. This is may be partly because of the absence of a recognized treatment for the disease, however, the prevalence and nature differ from country to country which may influence human behavioural responses.

**Aim:** This study aimed to investigated the beharioural response of the community towards self-medication practices during this COVID-19 pandemic and lockdown.

**Methods:** A cross sectional household and online survey was conducted during the months of June-to-August. The study was conducted among adult between age 18 above in communities of western Uganda who consented to participate in the study. Study participants were selected using a convenience sampling technique and sampling was done by sending a structured online questionnaire via Google forms and a printed copies questionnaire made available to other participants that did not use the online questionnaire

**Results:** The percentage of respondents that know about self-medication is (97%) and those that practice self-medication are approximately (88%). 97% of respondents have heard about self-medication either through health workers, media, family members, friends and/or school while 3% said they have not heard about self-medication. The percentage of respondents who practiced self-medication during COVID-19 pandemic is 57% while those that did not is 43%. There is statistically difference in the number of those that practice self-medication and those that do not p < 0.005 at 95% confidence interval. Also there was a statistically significant decrease in the number of respondents that practice self-medication during COVID-19 pandemic lockdown compare to the practice before the pandemic lockdown p < 0.05 at 95% confidence interval.

**Conclusion:** Our investigation showed adequate knowledge of self-medication and high level of self-medication practice with a decrease in self-medication practices during the COVID-19 pandemic lockdown compared to the practice before the lockdown.

## INTRODUCTION

The outbreak of coronavirus have prompted many countries around the world including Uganda to introduce lockdown as advised by WHO because of growing cases. In most countries so far, partial lockdowns as well as social distancing guidelines, frequent washing of hands and/or sanitizing have been put in place to curb the spread of the virus. Some of these restrictions may remain in place probably until a vaccine for the virus has been developed. In this regards many human behavioural patterns to life have been seriously affected raging from education, economic, health, social, religion etc.

In Uganda for example, total ban on people’s movement from one district to another, border closure, curfew and others are measures that have been employed to prevent the spread of the virus, however, many patients suffering from other diseases have little or no access to health care service as a result of restrictions especially on movement. Some studies have revealed significant changes in the way people live their lives during this lockdown which included avoiding crowded public places, wearing a face mask, avoiding physical contact, improving personal hygiene (e.g. frequent washing of hands and using hand sanitizer), changes in sleep and use of sleep substances, diet and physical activities (Bruine de Bruin & Bennett, 2020, Arora & Grey, 2020 and WHO, 2018). Some of these may have some behaviours and mental health implications

In the period of this pandemic, the way individual behaves is important to note because a change in the public behavior is an alternative to controlling outbreaks when there is no suitable pharmacological interventions. Furthermore, the behavioural patterns of an individual can influence their family, communities, society and social networks (Chen et al., 2017). Epidermics had been reported in a study which used mathematical models to affect people’s fears, emotions and ultimately their behavior (Bi et al., 2019). With respect to these behavioural changes resulting from the lockdown and restrictions, we envisaged that self-medication practices may be influenced by the way people try to cope with their health management at this crucial time.

Self-medication can be referred to as the intake of drugs, herbs or home remedies either on one’s own initiative, or on the advice of another person for physical or psychological ailments (Hernandez-Juyol and Job-Quesada, 2002). Common sources of self-medications include previous prescribed drug, pharmacist, family, friends, neighbours, internet, and suggestions from an advertisement etc. According to Osemene and Lamikanra, (2012), self-medication is a serious public health problem globally posing risks such as drug resistance, organ damage and deaths (2.9 – 3.7%) in the world because of drug-drug interaction.

The probability of drug abuse has been reported to be on the rise as a result of global increase in self-medication (McCabe et al., 2005, WHO 1998). Signs and symptoms of underlying disease conditions can be concealed by self-medication, therefore causing drug resistance, delay diagnoses and making the problem complex or difficult to deal with (Bauchner and Wise 2000, Calabresi and Cupini, 2005).

Although, several countries and cultures presents the nature and prevalence of self-medication differently globally, nevertheless the practices are mostly based on certain advantage or disadvantage obtained from past experiences with similar drugs (McCabe et al., 2005). The effects of self-medication can be positive when practiced correctly; such as saving scarce medical resources from being wasted on minor conditions, reducing the pressure on medical services where health care personnel are insufficient, lowering the costs of community funded health care program and reducing absenteeism from work due to minor symptoms (WHO, 2000). However it also has potential risks at individual level such as incorrect self-diagnosis and choice of therapy, incorrect route of administration, severe adverse effects, failure to recognize that the same active substance is already being taken under a different name and failure to recognize contraindications, interactions and precautions, incorrect route of administration, inadequate and excessive dosage and more. Also, self-medication could lead to high drug induced disease and public expenditure wastefulness.

Self-medication causes adverse reaction and prolonged suffering as well as other related problems such as wastage of resources and high resistance of pathogens. It is important to note that a current world-wide problem especially in developing countries where antibiotics are available without any prescription is antimicrobial resistance (Bennadi, 2014). The overall self-medication rate in sub-Sahara Africa varies from 11.9% to 75.7%. In Uganda, a high prevalence of self-medication was previously reported in the northern and southwestern region (Mouankie et al., 2011, Ocan et al., 2014, Lukyamuzi et al., 2020). Therefore, this research sought to investigated the beharioural response of the community towards self-medication practices during this COVID-19 pandemic and lockdown.

## MATERIALS AND METHODS

### Study design, site and population

A cross sectional household and online survey using traditional method (printed questionnaires) and information technology (Google form) was conducted during the months of June-to-August 2020 in western Uganda. The study was conducted among adult between age 18 above in communities of western Uganda.

### Sample size determination

The population of the western region, one of the four regions of Uganda was 8,874,862 according to 2014 census (Uganda Bureau of Statistics 2013, *Citypopulation.de* 2016). With about a population growth rate of 3.32% (https://worldpopulationreview.com/countries/uganda-population), the sample size of the study was calculated using the OpenEpi sample size calculator (Open source statistics for public health. (2013). Accessed: March 9, 2020: http://openepi.com/SampleSize/SSPropor.htm). The estimated sample size was 280, with a confidence level of 90% using an anticipated frequency (p) of 50%.

Study participants were selected using a convenience sampling technique and sampling was done by sending a structured online questionnaire via Google forms to some participants. Also a printed hard copied self-administered questionnaires were made available to other participants that did not use the online questionnaire. The questionnaire consisted of 20 questions covering two parts. The first part of the questionnaire included demographics data of the participants (e.g. age, gender, education, occupation). While the second part of the questionnaire focused on the self-medication practice before and during COVID-19 pandemic. To ensure anonymity and confidentiality, no names or email addresses were asked. Those who participated willingly consent to take part in the study and for quality control, strict parameters were set to ensure participants answer the questionnaire once.

### Data management

The researcher checked all filled data collection tools for validity and completeness. Any discrepancies in the entries were corrected by referring to the source documents (questionnaires).

### Statistical analysis

The data were entered into MS-Excel and OpenEpi version 3.01. Odd ratio and 95% Confidence interval were calculated using MS-Excel, two-sample independent t test, ANOVA and Chi-square were the tools used for data analysis with statistical significance level of 0.05.

### Ethical Consideration

Expedited ethical clearance was sought from Kampala International University Ethical Review Committee and registered as Nr.UG-REC-023/201914. Consent to participate was acquired through online submission of questionnaires and traditional printed and filled questionnaires by the participants.

## RESULTS

### Sociodemographic Characteristics of Participants

Our study showed the sociodemographic features of the respondents where most of them were males 149, (54.8%) and females 124, (45.2%) between ages 25 -34 years (43.8%), Christian (85.7.8%) and others as shown in Table 1. The percentage of respondents that know about self-medication is (97%) and those that practice self-medication are approximately (88%). Previous studies report that generally, people are aware of the effects, potential side effects, misuse and abuse of OTC drugs.10-12. Our result showed that 97% of respondents have heard about self-medication either through health workers, media, family members, friends and/or school while 3% said they have not heard about self-medication. There is a statistically significant difference between those that have heard of self-medication and those that have not. The number of respondents that knew about self-medication were 146 (98%) male and 119 (96%) female while those that are not aware of self-medication were 3 (2%) male and 5 (4%) female (OR = 2.0, 95%CI = 0.49, 8.73)

**Table 1:** Socio-demographic characteristics of the study respondents (n = 272)

|  |  |  |  |
| --- | --- | --- | --- |
| Characteristics |  | Frequency (n) | Percentage (%) |
| Gender | Male | 149 | 54.8 |
|  | Female | 123 | 45.2 |
| Age | 18 - 24 | 90 | 33.0 |
|  | 25 - 34 | 119 | 43.8 |
|  | 35 - 54 | 51 | 18.8 |
|  | 55 and above | 12 | 4.4 |
| Education status | Uneducated | 10 | 3.7 |
|  | Primary School | 19 | 7.0 |
|  | Secondary school | 11 | 4.0 |
|  | Certificate/ Diploma | 137 | 50.4 |
|  | Bachelors/ Masters/PhD | 95 | 34.9 |
| Occupation | Unemployed | 19 | 7.0 |
|  | Student | 70 | 25.7 |
|  | Lecturer/Teacher | 48 | 17.6 |
|  | Health worker | 38 | 14.0 |
|  | Merchant/Peasant | 80 | 29.4 |
|  | Others | 17 | 6.3 |
| Religion | Christianity | 233 | 85.7 |
|  | Islam | 36 | 13.2 |
|  | Others | 3 | 1.1 |

### Self-medication Practices Before and During COVID-19 Pandemic

Among the 272 respondents, those who had practice self-medication before COVID-19 pandemic were 239, (88%) and those that have not were 33, (12%). Those that have practice self-medication were statistically significantly more that those who have not (P < 0.005 at 95% Confidence interval) (Figure 2). During COVID-19 pandemic, the respondents who practiced self-medication were 156, (57%) while those that did not were 116, (43%). There is statistically significant difference in the number of those that practice self-medication and those that do not p < 0.005 at 95% confidence interval (Figure 3). Also there was a statistically significant decrease in the number of respondents that practice self-medication during COVID-19 pandemic lockdown compare to the practice before the pandemic lockdown p < 0.05 at 95% confidence interval (OR = 5.39, 95% CI = 3.48, 8.32).

**Figure 1:**
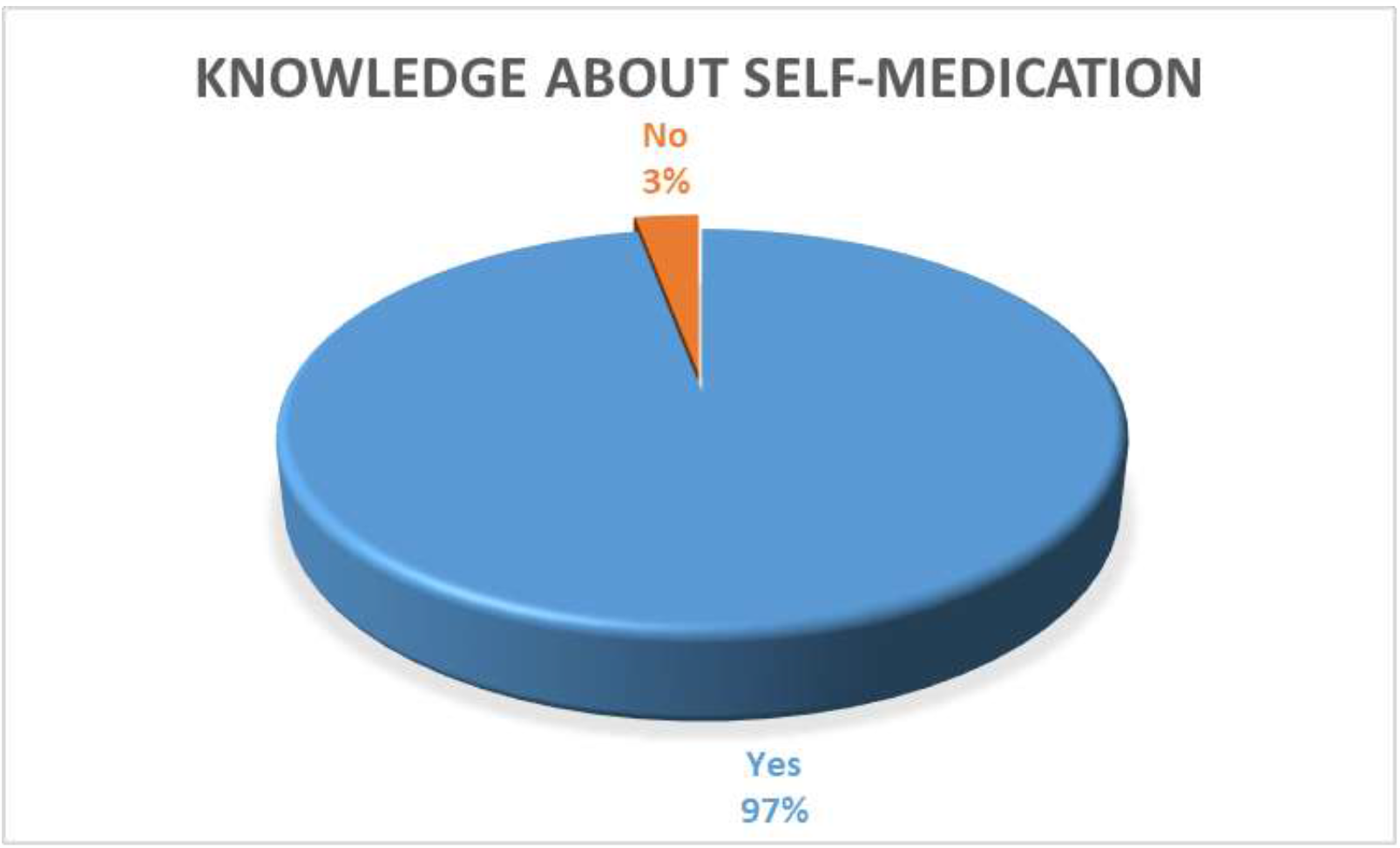
Pie chart showing the awareness of the respondents on self-medication

**Figure 2:**
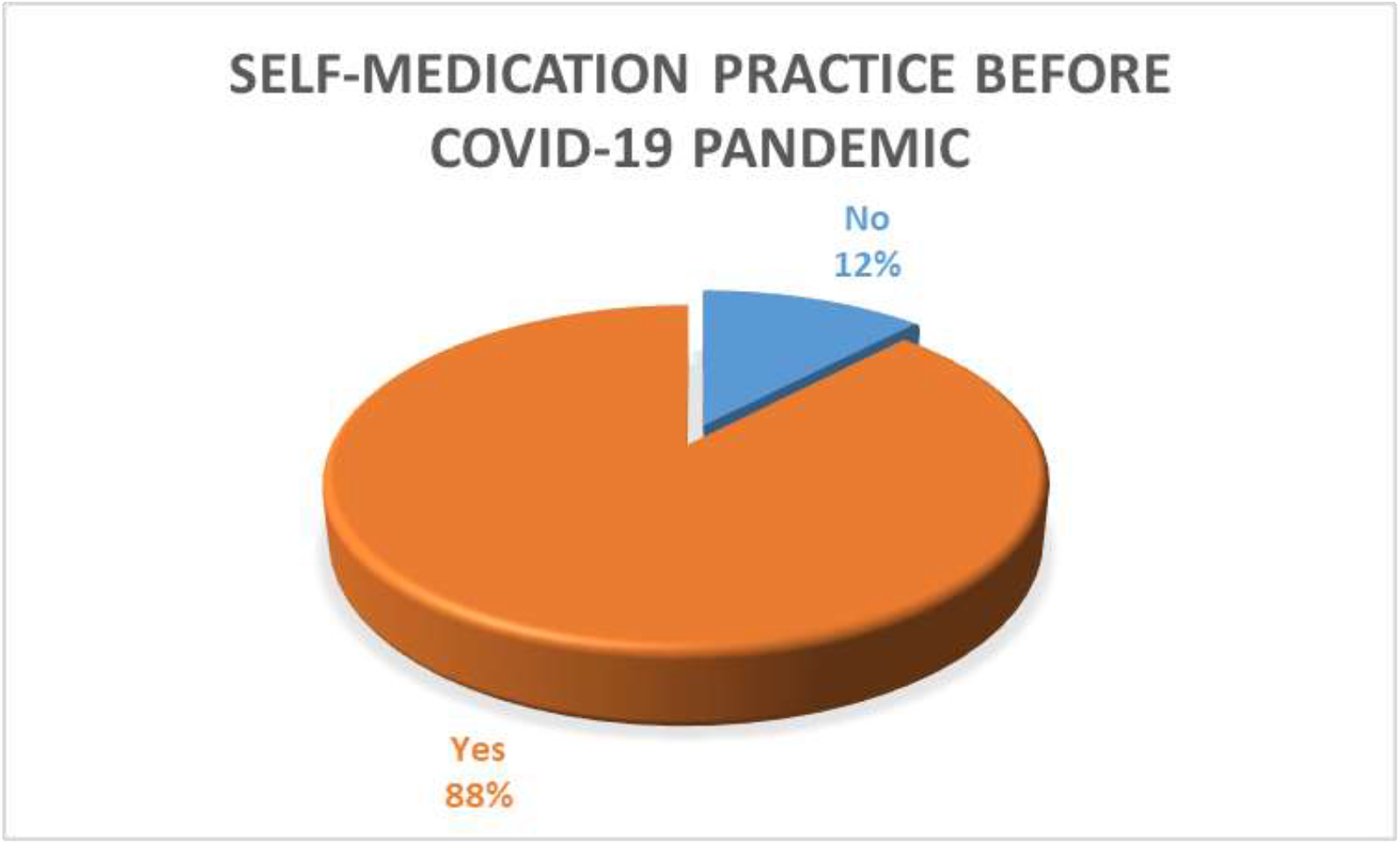
Pie chart showing self-medication practice before COVID-19 pandemic.

**Figure 3:**
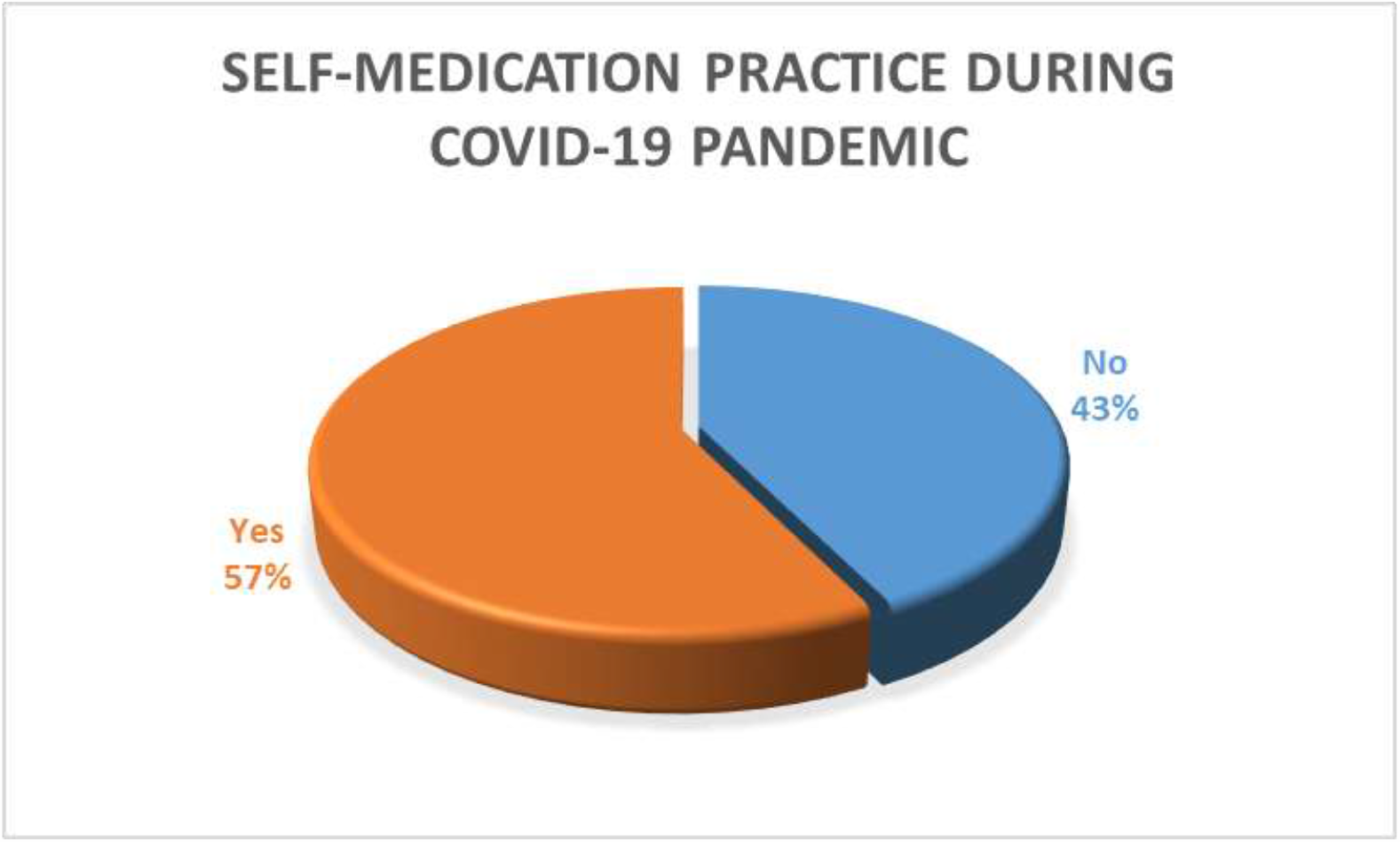
Pie chart showing the self-medication practice during COVID-19 pandemic

### Frequency of Self-medication Before and During COVID-19 Pandemic

Considering how often the respondents practice self-medication, our results showed that majority of the respondents sometimes practice (68, 46% male and 64, 52.5% female) self-medication before COVID-19 pandemic lockdown. 19 (12.8%) male and 13 (10.7%) female never practice self-medication before the lockdown. The male respondents that always practice self-medication were 12 (8.1%) and female (16) 13.1% while those that rarely practice self-medication are 49 (33.1%) male and (29) 23.8% female (OR = 1.0, 95%CI = 1.31, 5.65). A Chi-square test shows a statistical significant difference between male and female p < 0.05 (Figure 4). During the COVID- 19 lockdown, the respondents who sometimes practice self-medication were 42 (28.4%) in male and 35 (28.7%) in female. Those respondents who never engage in self-medication at all are 57 (38.5%) in male and 51 (41.8%) in female. 42 (28.4%) male and 35 (28.7%) female sometimes practice self-medication while 42 (28.4%) male and 21 (17.2%) female rarely practice self-medication during the pandemic (OR = 0.96, 95%CI = 1.6, 4.2). A Chi-square test shows a statistical significant difference between male and female p < 0.05 (Figure 5).

**Figure 4:**
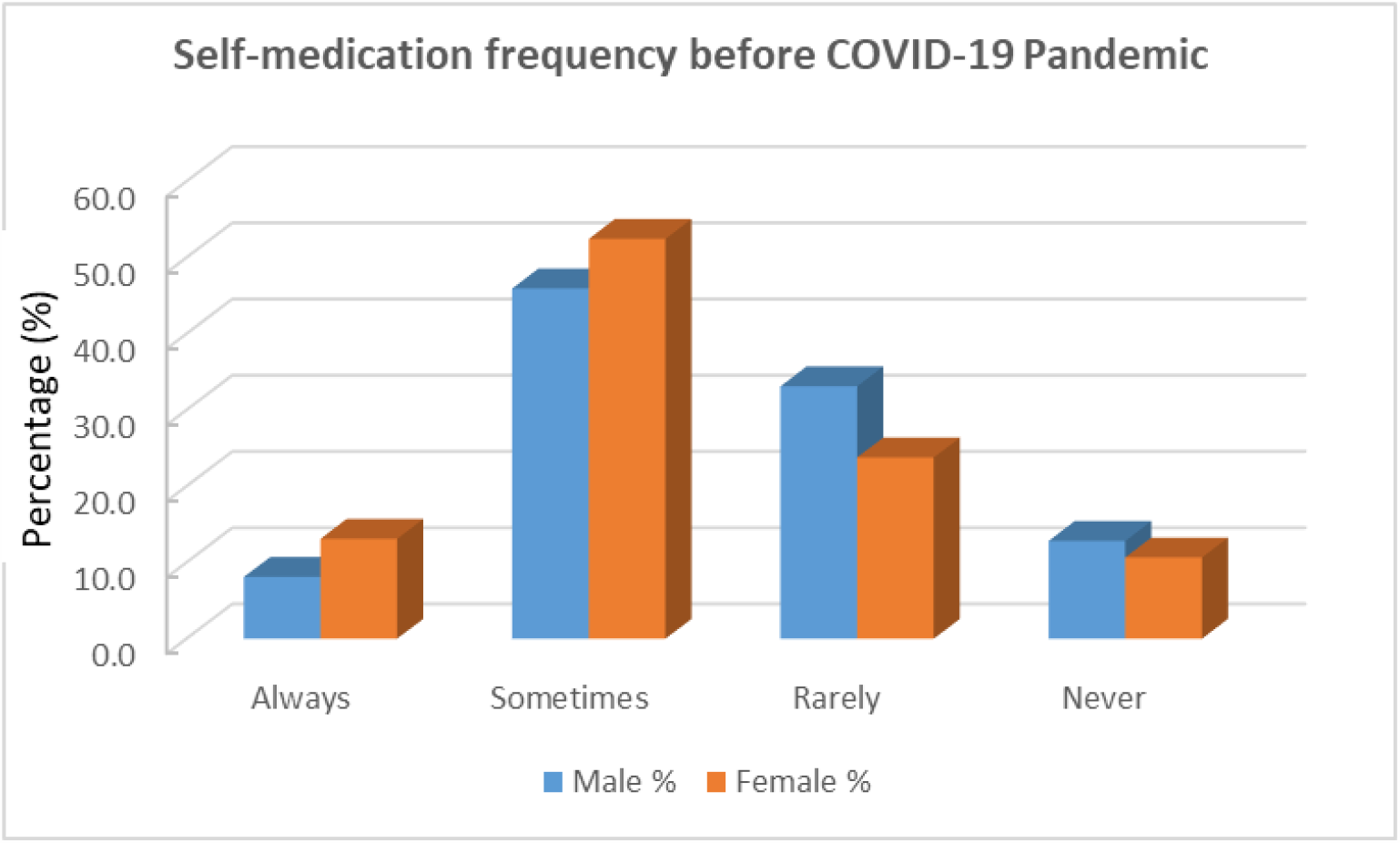
Showing the frequency of self-medication before COVID-19 pandemic. A Chi-square test shows a statistical significant difference (P = 0.0000000081) between male and female respondents.

**Figure 5:**
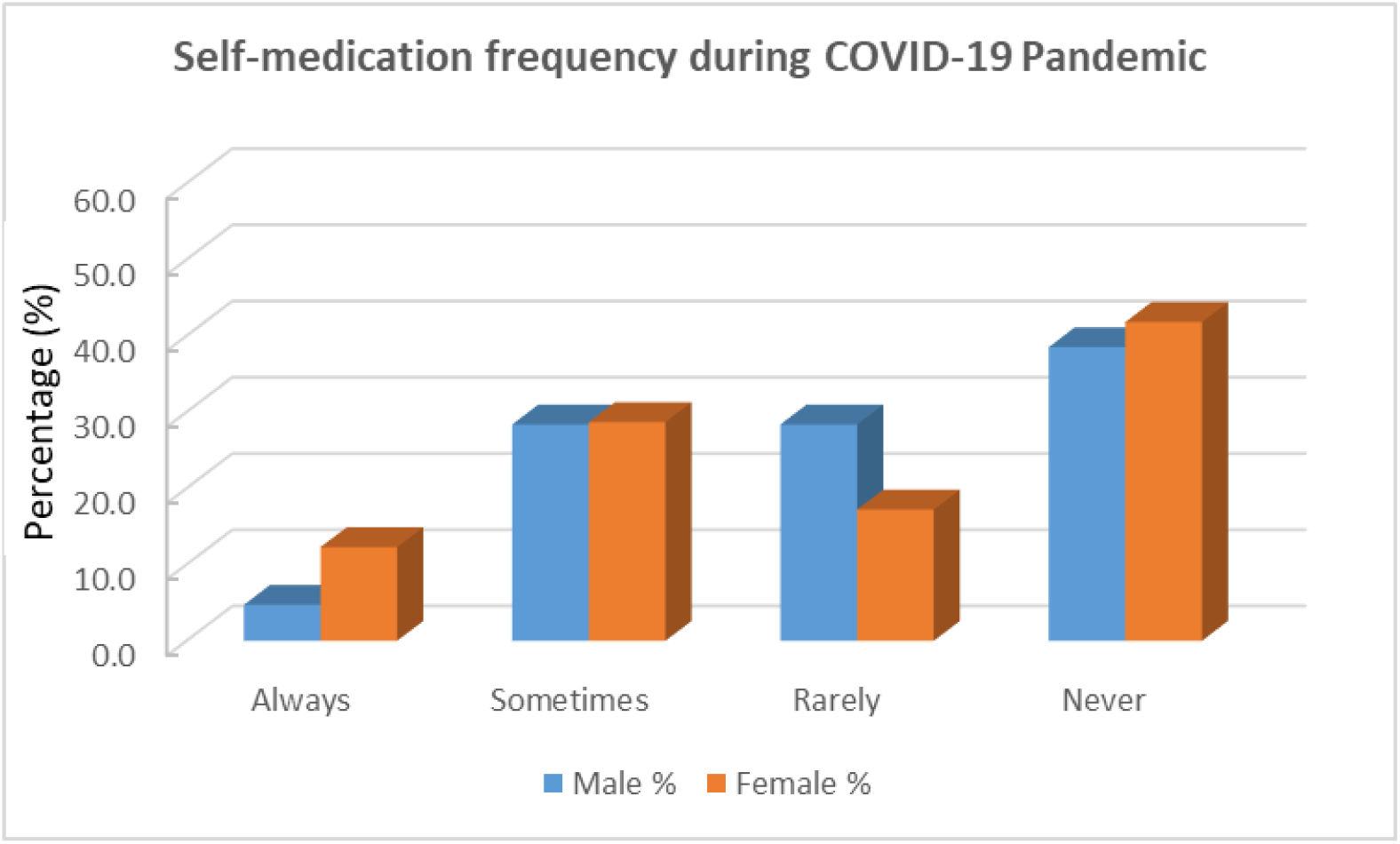
Showing the frequency of self-medication during COVID-19 pandemic. Chi-square test shows a statistical significant difference (P = 0.0000000081) between male and female respondents.

### Incidence of Sicknesses Before and During COVID-19 Pandemic

Among 272 respondents, those that never fall sick before COVID-19 pandemic lockdown is 17 (7.4%) in which increases to 81 (35.2%) during the lockdown. 7 (3%) never, 86 (37.2%) sometimes, 121 (52.4%) rarely practice self-medication before COVID-19 pandemic lockdown, however, 4 (1.7%) never, 58 (25.2%) sometimes, 87 (37.8%) rarely practice self-medication during the lockdown (Figure 6). Our result revealed a statistical significant decrease (P < 0.05) in the incidence of sickness before and during COVID-19 pandemic lockdown.

**Figure 6:**
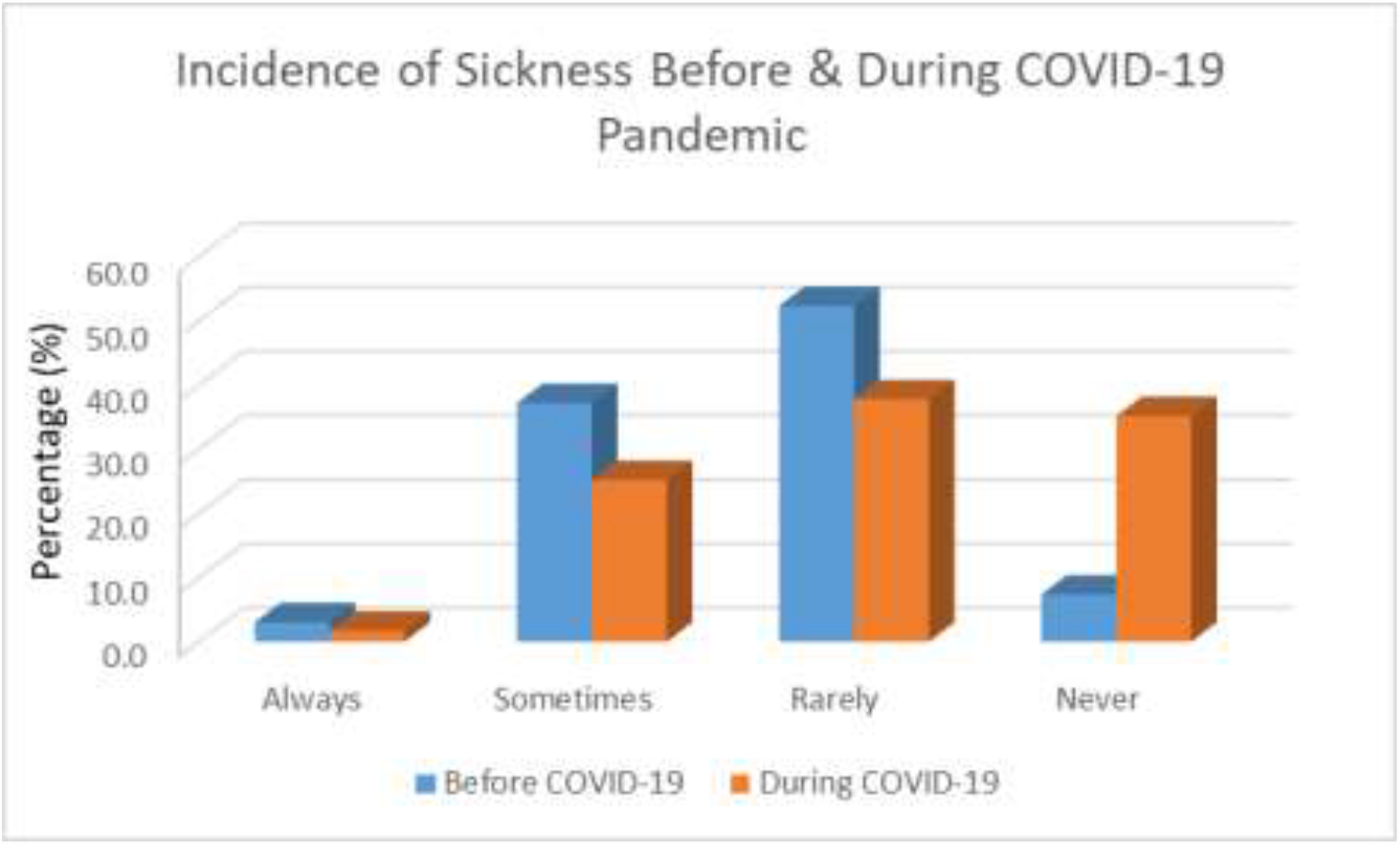
Showing the incidence of sickness before and during COVID-19 pandemic

### Reason for Self-medication Practice

The reasons people self-medicate are numerous, some of which were investigated in this study and from our respondents 7% fear visiting health facility or hospital, 9% fear been diagnosed COVID-19 positive, 15% lack the means to get to the health facility/hospital, 37% consider self-medication affordable while 32% said it is convenience (Figure 7). Generally, most of the respondents (41%) opined that self-medication should be discouraged, 21% said it should be encourages, 18% said it is time saving and 23% said it is cheap (Figure 8).

**Figure 7:**
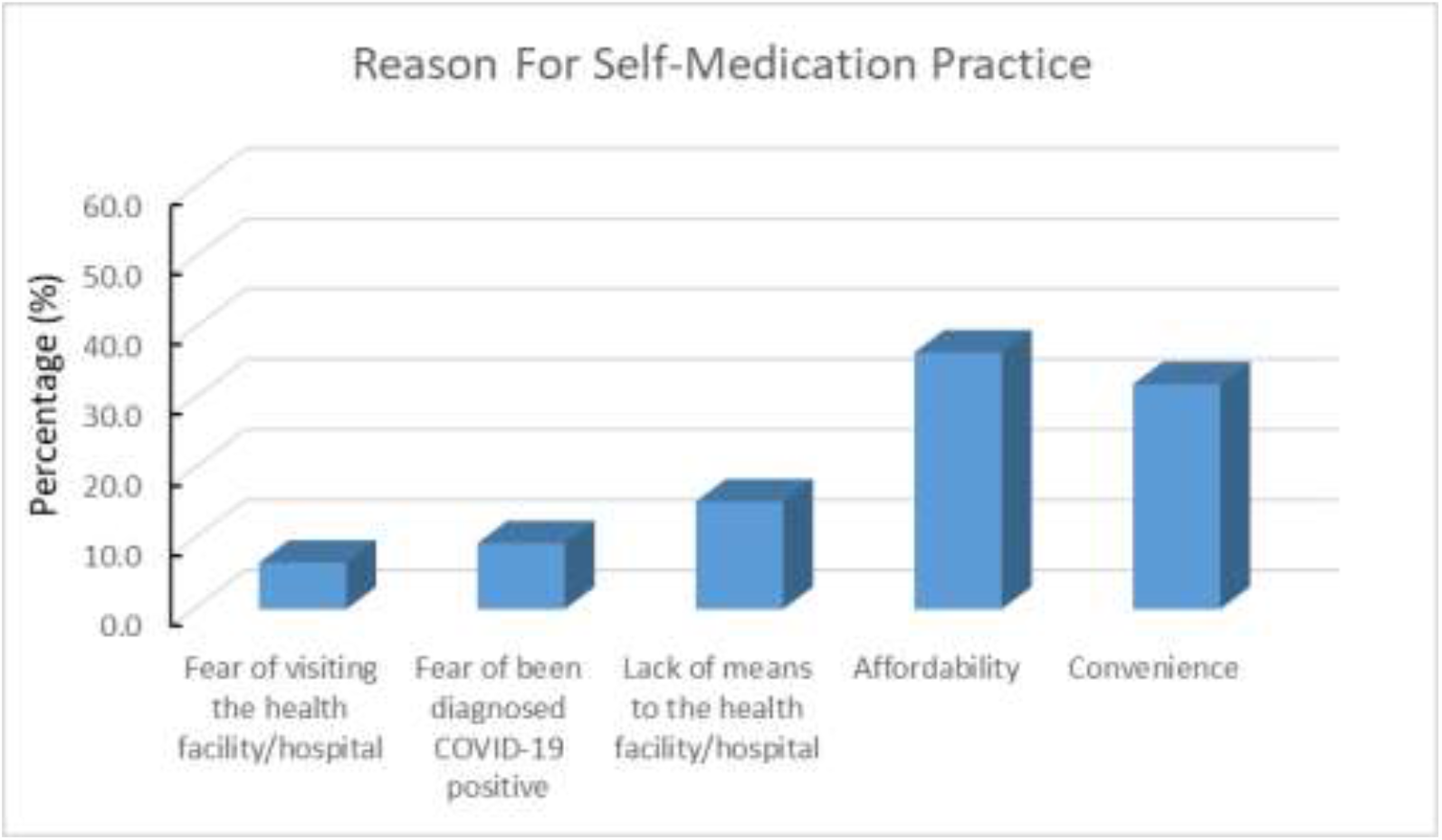
Showing reasons for self-medication practice

**Figure 8:**
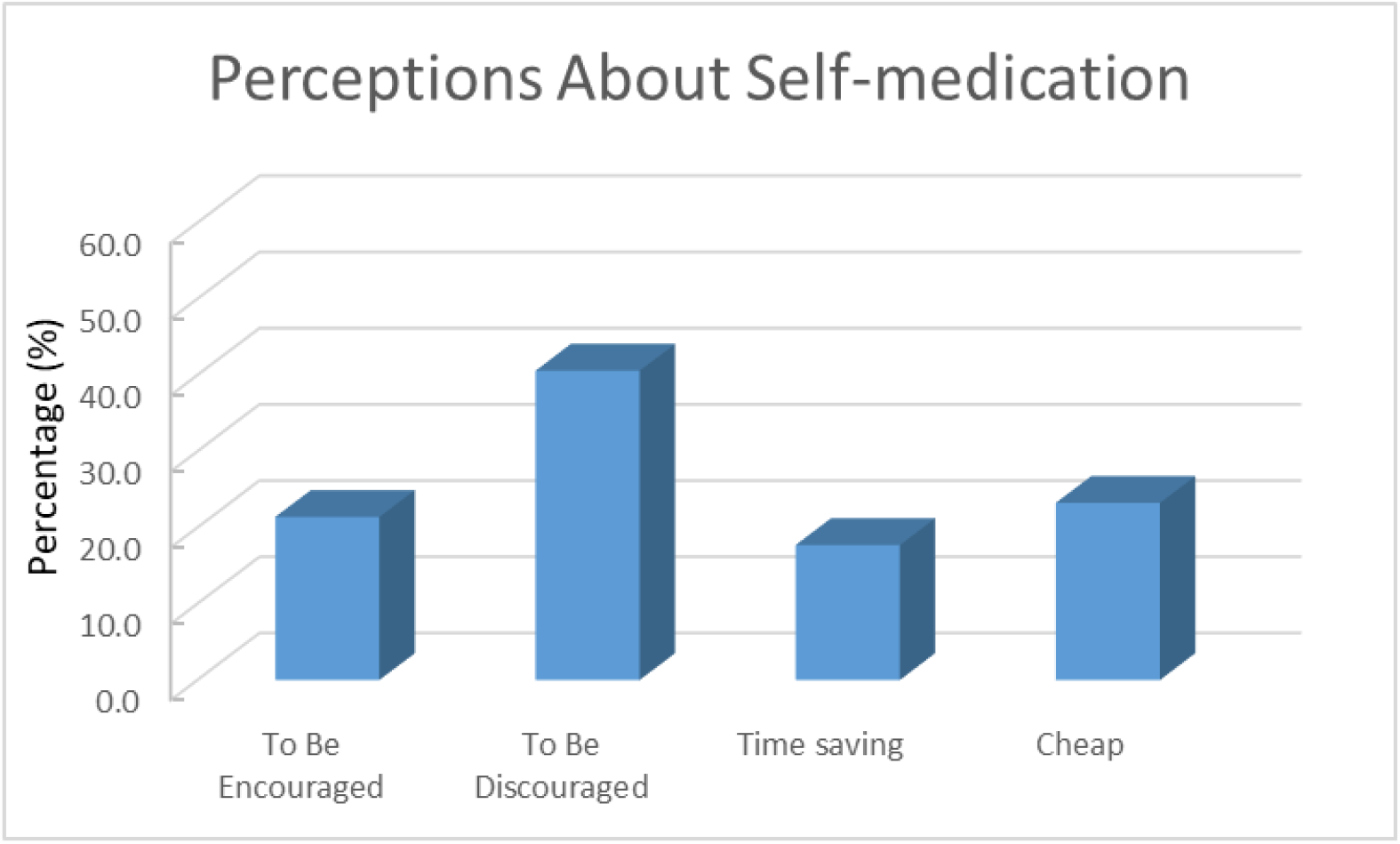
Showing the perceptions of respondents on self-medication

## DISCUSSION

Many factors have been reported to affect self-medication practice which includes availability of drugs, education society and many more (Vizhi & Senapathi 2010; Montastruc et al., 1997; Habeeb et al., 1993). Our investigation revealed that most of the respondents know about self-medication and practice it (Figure 1). The rate of self-medication was high at 88% before COVID- 19 lockdown with more practice shown in the female, but was reduced to 57% during the lockdown (Figure 2 & 3). The higher rate of self-medication found among females compare to males in this study is consistent with previous studies (Figueiras et al., 2000 and Awad et al., 2005)

The World Health Organization (WHO) has continue to promote self-medication as a means to treat common diseases in the community considering it to be beneficial and helping in decreasing the burden on health care services especially in developing countries (WHO, 1986). In this respect self-medication when practiced correctly could be a great benefit to the community during the COVID-19 lockdown as it allows patients to become responsible and build confidence to manage their health. Nevertheless, previous studies has showed quite a number of negative effects from substance use resulting from social isolation such as COVID-19 lockdown and restrictions, with probable drug abuse associated more to socially isolated people (Hawkley and Cacioppo, 2010; Dyal and Valente, 2015; Nino et al., 2016), and most likely diagnosed with substance use disorder (Chou et al., 2011).

Our result shows a significant decrease in the percentage of people that practiced self-medication before COVID-19 pandemic compared to the percentage of people that practiced self-medication during COVID-19 pandemic lockdown (Fig. 4 & 5). Also there was a decrease in the incidence of sickness during COVID-19 lockdown as many of the respondents never fall sick during lockdown (Figure 6). The decrease in the incidence of sickness may be due to the change in behavior of people as a result of the measures put in place to combat the spread of the disease. During the lockdown, there is a high propensity for people pay more attention to their health as people were encouraged to practice more personal hygiene such as washing of hands regularly and/or using sanitizers, wearing face masks and maintaining social distance which may have resulted in the improvement of people’s health status in the community. This is consistent with the report that the incidence of diseases such as infections, trachoma, diarrhea-related diseases, pneumonia and many more can be decreased by maintaining good personal hygiene (WHO, 2018).

Major reason for self-medication were convenience, affordability, lack of means to the health facility/hospital, fear of been diagnosed COVIC-19 positive and fear of visiting the health facility/hospital, time saving (Figure 7). On perception of respondents to self-medication, most of them (40.6%) opined that self-medication should be discouraged (Figure 8), however the percentage of respondents practicing self-medication is still high. This may have emanated from the fact that self-medication practice reduces the time spent waiting for a doctor, can save life in emergency states and affordable cost of health care (Almasdy & Sherrif, 2011). Also, the World Health Organization recommended that self-medication can assist in prevention and treatment of sicknesses where consulting a doctor is not necessary, therefore making treatment of common illness cheaper. Nevertheless, inappropriate use of medication could lead to a rise in diseases triggered by abuse and cause waste of public funds (WHO, 2000).

## CONCLUSION

Our investigation showed that majority of populace have adequate knowledge of self-medication as evidenced by higher percentage of respondents in this category and there was a high level self-medication practice prior to COVID-19 pandemic lockdown. However, there was a decrease in self-medication practices during the COVID-19 pandemic lockdown compared to the practice before the lockdown. This decrease correlates with reduction in the incidence of sickness during the lockdown which could have resulted from practice of good personal hygiene as recommended and campaigned by WHO in order to prevent the spread of the COVID-19. However, WHO has recommended that self-medication should be taught with the right approach and also properly regulated so as to prevent drug resistance problems associated with the wrong use of drugs globally and particularly in developing countries where there is no control of the use antibiotics.

## Data Availability

The metadata used to support the findings of this study are available from the corresponding author upon request.

